# Deep learning based phenotyping of medical images improves power for gene discovery of complex disease

**DOI:** 10.1101/2023.03.07.23286909

**Authors:** Brianna I. Flynn, Emily M. Javan, Eugenia Lin, Zoe Trutner, Karl Koenig, Kenoma O. Anighoro, Eucharist Kun, Alaukik Gupta, Tarjinder Singh, Prakash Jayakumar, Vagheesh M. Narasimhan

## Abstract

Electronic health records (EHRs) are often incomplete and inaccurate, reducing the power of genome-wide association studies (GWAS). Moreover, the variables within these records are often represented in binary codes, masking variation in disease severity among individuals. For some diseases, such as knee osteoarthritis (OA), radiographic assessment is the primary means of diagnosis and can be performed directly from medical images. In this work, we trained a deep learning model (DL-binary) to ascertain knee OA cases from anteroposterior (AP) dual-energy absorptiometry (DXA) scans and achieved clinician level performance. Applying this model across 29,257 individuals from the UK Biobank (UKB), we identified 2,603 (240%) more cases than currently diagnosed in the ICD-10 record. Individuals diagnosed as cases by DL-binary had higher rates of self-reported knee pain, knee pain for longer durations and with increased severity compared to control individuals. We trained another deep learning model to measure the minimum knee joint space width (mJSW), a quantitative phenotype linked to knee OA severity. Despite the DL-binary phenotype and mJSW being highly genetically correlated (92%), the heritability of mJSW was an order of magnitude greater than the ICD-10 code M17 or DL-binary phenotypes. In a GWAS run on mJSW, we identified 18 genome-wide significant loci, as opposed to 1 and 6 at the same sample size using either case-control (DL-binary and ICD-10 code M17) phenotype. This improved power also translated to better polygenic risk score (PRS) prediction for knee OA diagnosis in a holdout dataset of 371,686 individuals. We also show that reduced mJSW, but neither case-control phenotype is associated with increased risk of adult fractures, a leading cause of injury-related death in older individuals. For diseases with radiographic diagnosis, our results demonstrate the enormous potential for using deep learning to phenotype at biobank scale, both for improving power for gene discovery and for epidemiological analysis.

## Introduction

For most complex disease traits, clinical endpoints are usually binary (case-control) in nature. In particular, data on disease outcomes from population scale biobanks are only available through recorded ICD-10 billing codes or self-reported diagnosis^1–3^. While these datasets have provided invaluable insights into the genetic basis of disease, case ascertainment based solely on information available in the EHR or from self-reports can be biased by a multitude of factors including differences in how patients were billed^4^, differential diagnosis due to assessment by clinicians (non-specialist vs specialist)^5^, or differences in classification or diagnosis based on disease severity^6^.

An alternate approach to ascertaining disease status might be to directly perform clinical-grade assessment from a patient’s medical images using a consistent diagnosis protocol. However, this is difficult to achieve at biobank scale where sample sizes can range from hundreds of thousands if not millions of individuals^1^. Importantly, for musculoskeletal diseases such as knee OA, radiography is the routine course of diagnosis in the clinic as well as to assess important markers associated with disease progression such as sclerosis, osteophytosis (bone spurs) and narrowing of the space between the femur and tibia, also known as the knee joint space^7^. For such radiographically diagnosed diseases, computer vision approaches for automated phenotyping based on training data from clinicians offer the potential to ascertain both case status and disease severity at scale. Such approaches have already been used for determining pneumonia and SARS-CoV-2 cases from chest X-ray images, with reported accuracy even higher than expert radiologists based on ground truth from molecular information^8,9^.

Taking advantage of these developments in computer vision, recent genetic studies have successfully applied these methods to generate image derived phenotypes (IDPs) of distribution of body fat, heart structure, liver fat percentage, and brain morphology, and have linked these novel traits with genome-wide significant loci^10–14^. While some recent studies on musculoskeletal disease employ these novel phenotyping approaches^15–17^, neither these nor the studies on other traits have specifically investigated how generating quantitative IDPs that underlie binary disease status could be used to improve power for gene discovery at biobank scale.

Quantitative measurements which provide information about variation in the severity of progression of the disease are already routinely utilized in predicting an individual’s risk for complex disease in the clinic. For example, LDL cholesterol levels are a quantitative biomarker measured in blood samples, and are used as a primary biomarker to assess risk for myocardial infarction, among the leading causes of death worldwide^18^. Multiple lines of functional evidence suggest that LDL cholesterol levels are also causally linked to heart disease and lowering LDL levels over an entire lifetime through the use of statins is the most widely used long term prescription medication^19^. In theoretical work, it has been demonstrated that with equal sample size and when the proportion of cases in a case-control design is equivalent to the prevalence of the disease in the population, the power of a case-control association study is considerably lower than that of a quantitative association study. This is in part because key information about variation in the trait in the sample population is lost when transforming a continuous trait into a binary one^20^.

Building on these foundational ideas, in this work we first trained a binary classification model to identify knee OA cases at clinical level performance and deployed this at biobank scale to compare our radiographically obtained results to the ICD-10 record. Second, we trained an image segmentation algorithm to obtain a quantitative measurement highly correlated with knee OA severity, mJSW, to examine differences in power between GWAS carried out using quantitative approaches versus a case-control design. Third, we generate PRS for each phenotype to evaluate if improvements in statistical power to find novel loci translate to better prediction of ICD-10 record knee OA (M17) in a hold-out dataset of over 300,000 individuals. Finally, we examined epidemiological associations to link our IDPs with an outcome of major clinical relevance.

## Results

### Dataset, and quality control of DXA imaging and genetic data

To study the genetic basis of knee phenotypes, we jointly analyzed paired DXA imaging and imputed genome sequence data of 42,284 individuals in the UKB. We first restricted the dataset to individuals of white British ancestry, applied standard variant and sample QC and analyzed 12.1 million common bi-allelic SNPs with minor allele frequency > 0.1%^1^ (**Methods**: Genetic QC). Next, as the bulk imaging data from the UKB comprised of DXA images that reflect scans of different body parts, we used a deep learning approach^15^ to subset the imaging dataset to only AP view knee scans. We then removed individuals that had outlier image resolutions or poor quality DXA scans, and padded images to a standard size for processing (see **Methods**: Image segmentation, phenotype measurement and quality control). Post quality-control, we were left with combined imaging and genetic data for a total of 29,257 individuals aged between 46 to 81 with a median age of 64, and a sex ratio of 0.99, consistent with the overall distribution in the UKB (**Methods**: UKB participants and dataset).

### Automated phenotyping of knee OA achieves clinician level performance

To perform automated phenotyping for knee OA based on radiography, we used a binary classification approach based on the Kellgren-Lawrence (KL) grading system^21^ (usually graded 0-4, where a 4 is considered the most severe case of radiographic OA) to determine case or control status for each individual reflecting different levels of joint space narrowing, subchondral sclerosis, and the presence of osteophytes. Cases were considered individuals with a KL grade of 3 or higher - severe enough that annotating clinicians would consider a candidate for joint replacement surgery in the clinic. Controls were considered individuals who would not be candidates for joint replacement - a grade 2 or lower (**Methods:** Binary classification: DXA scan annotation procedure). To train the deep learning model, we obtained case-control assessment on 546 images based on the annotations of three board-certified orthopedic surgeons who independently assessed each image. We then split the dataset so that 80% (436 images) of the data was used for training and 20% (110 images) was used for validation. We next trained a binary classifier (which we refer to as DL-binary) using transfer learning with the ResNet-101 architecture^22^ (**Methods**: Binary classification: Network architecture and model training). The sensitivity and specificity of our model on validation data (that is not used as part of the training process) was within the range of the sensitivity and specificity obtained between two clinicians grading the same set of images (Clinician sensitivity: 0.92 ± 0.05, DL-binary sensitivity: 0.82 ± 0.07 Clinician specificity: 0.95 ± 0.05, DL-binary specificity: 0.95 ± 0.06.) (**Fig. 1c, d**).

**Fig. 1:**
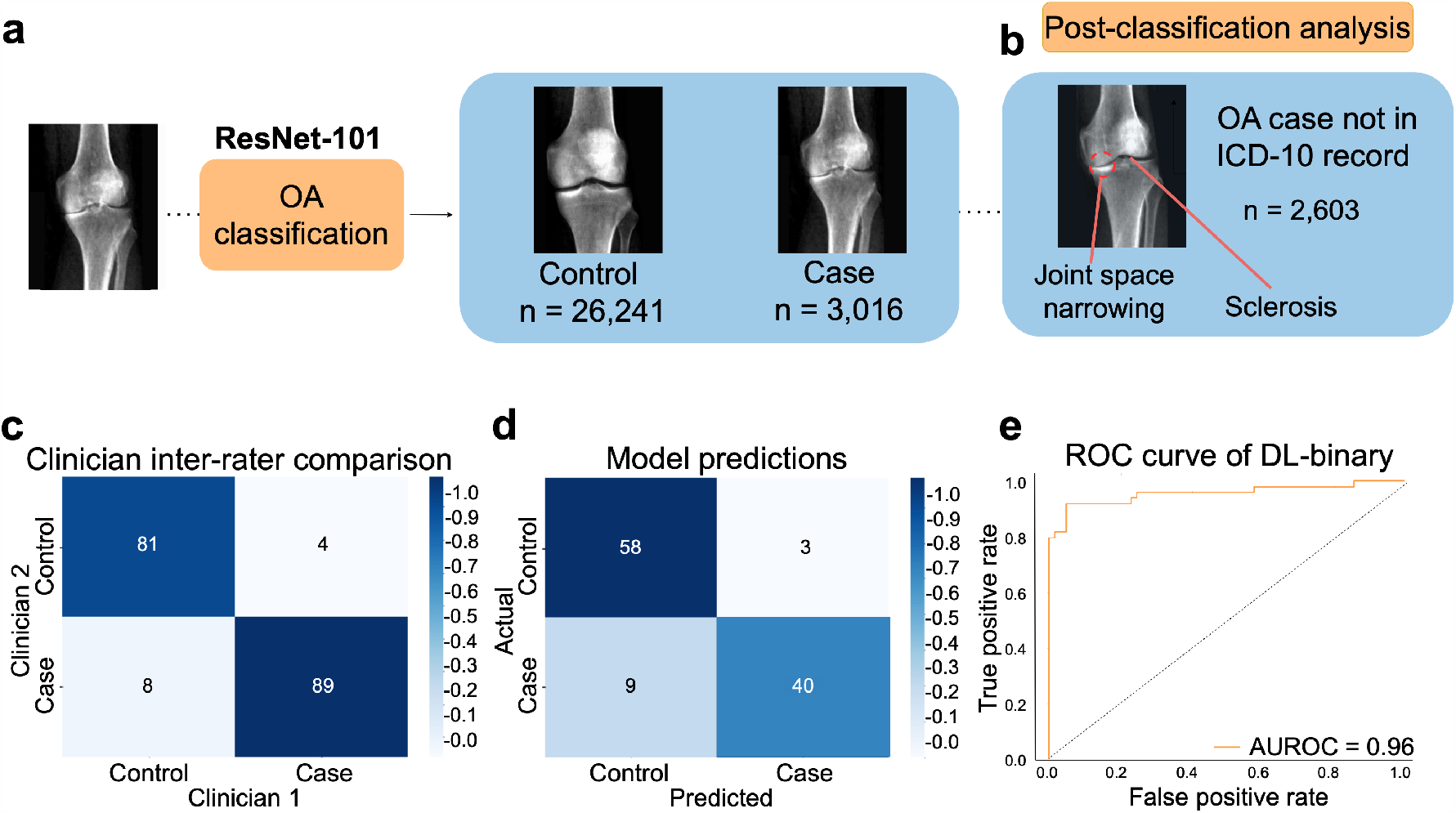
A deep learning process for automated phenotyping of radiographic knee OA. **a** ResNet-101 based classifier for binary classification of knee OA, showing an example of a typical individual diagnosed as a case compared to a control individual. **b** Post-classification analysis using highlighting regions of the knee that are discriminatory for knee OA. We confirmed joint space narrowing and sclerosis of the bone (important features for case classification) are present in cases not reported in the ICD-10 record but identified by the model. **c** Inter-rater comparison of two clinicians grading a total of 200 AP view knee DXA scans split roughly equally between cases and controls. **d** Confusion matrix showing performance of the DL-binary model on validation data. **e** Receiver operating characteristic (ROC) curve for DL-binary, showing performance of the model under different classification thresholds.

### Image based phenotyping reveals twofold more cases compared with ICD-10 records

We next deployed our trained model on the remaining 28,725 images of knee DXA scans from the dataset. We considered an individual a ‘case’ if our model predicted the individual to have knee OA on either the left or the right knee, and a control otherwise in line with the ICD-10 code M17 for knee OA. We then assessed how many cases were determined by our deep learning based binary classification of radiographic OA as compared to what already exists in the ICD-10 code M17. We found that after deploying the DL-binary classifier, we determined 2,603 more cases compared with the ICD-10 code for knee OA (ICD-10 code M17 1,085 cases, DL-binary 3,016 cases) (**Fig. 1a**). To provide additional support for cases reported by DL-binary that were not already reported in the ICD-10 code M17, our clinical team examined 100 individuals manually and confirmed the presence of osteophytes, reduced joint space and in some cases subchondral sclerosis (**Fig. 1b**). As these alone may not be diagnostic, we also investigated associations with three self-reported measures of knee pain in the UKB: knee pain experienced in the past month (binary), knee pain for 3+ months (binary, and reflecting knee pain experienced over a long duration) and rating of knee pain in the past three months (scale from 0 - 10). We found that in individuals who were newly identified as cases, the rate of self-reported knee pain was significantly higher compared to control individuals (individuals not diagnosed by ICD-10 code M17 or by DL-binary) across all three measures we examined (recent pain reported as knee pain in the past month: 49.4% in cases and 27.2% in controls, chi-square statistic = 536.6, p = < 2.2 × 10^−16^, chronic pain as determined by knee pain lasting 3 or more months: 80.4% in cases and 70.6% in controls, chi-square statistic = 28.29, p = 1.5 × 10^−7^ and severity of pain reported in the last 3 months: mean rating of 3.33 in cases and 2.58 in controls, t-test p = 1.45 × 10^−15^). These results suggest that knee OA is likely underdiagnosed in the ICD-10 record and that our approach is capable of identifying additional true cases not already present in the EHR.

### Image segmentation to measure joint space width

To examine knee OA severity beyond simple case-control assessment, we developed a method to obtain a quantitative measurement from knee DXA scans known to be highly associated with the disease: the minimum inter-bone joint space between the femur and tibia, which we refer to as mJSW. To perform automated measurement on the UKB dataset, we first collected training data for 63 DXA scan derived images of the knee (40 training, 23 validation). On each of these images we labeled the positions of the femur, tibia and fibula at pixel level, which were then validated by a team of clinicians. We then trained a deep learning model based on the U-Net architecture^23^ with a 34-layer ResNet encoder^22^ to perform semantic segmentation of the femur, tibia and fibula in each DXA image at pixel-level resolution (**Fig. 2a**). After quality control and image normalization (**Methods**: DXA scan image quality control and standardization), we computed mJSW by measuring the distance between the femur and tibia along multiple positions on the medial, lateral and center axes of the joint. We then computed the average of these distances for each leg (**Fig. 2b**). The mJSW measurement is defined as the smallest of the two averages for either leg, returning one phenotype measurement per individual. If the individual only had a right or left leg DXA scan, this was used as the mJSW measurement for that individual. To standardize mJSW measurements across image resolutions, we regressed each of the joint space lengths on the overall height of the individual (**Methods**: Image segmentation: Measurement and quality control).

**Fig. 2:**
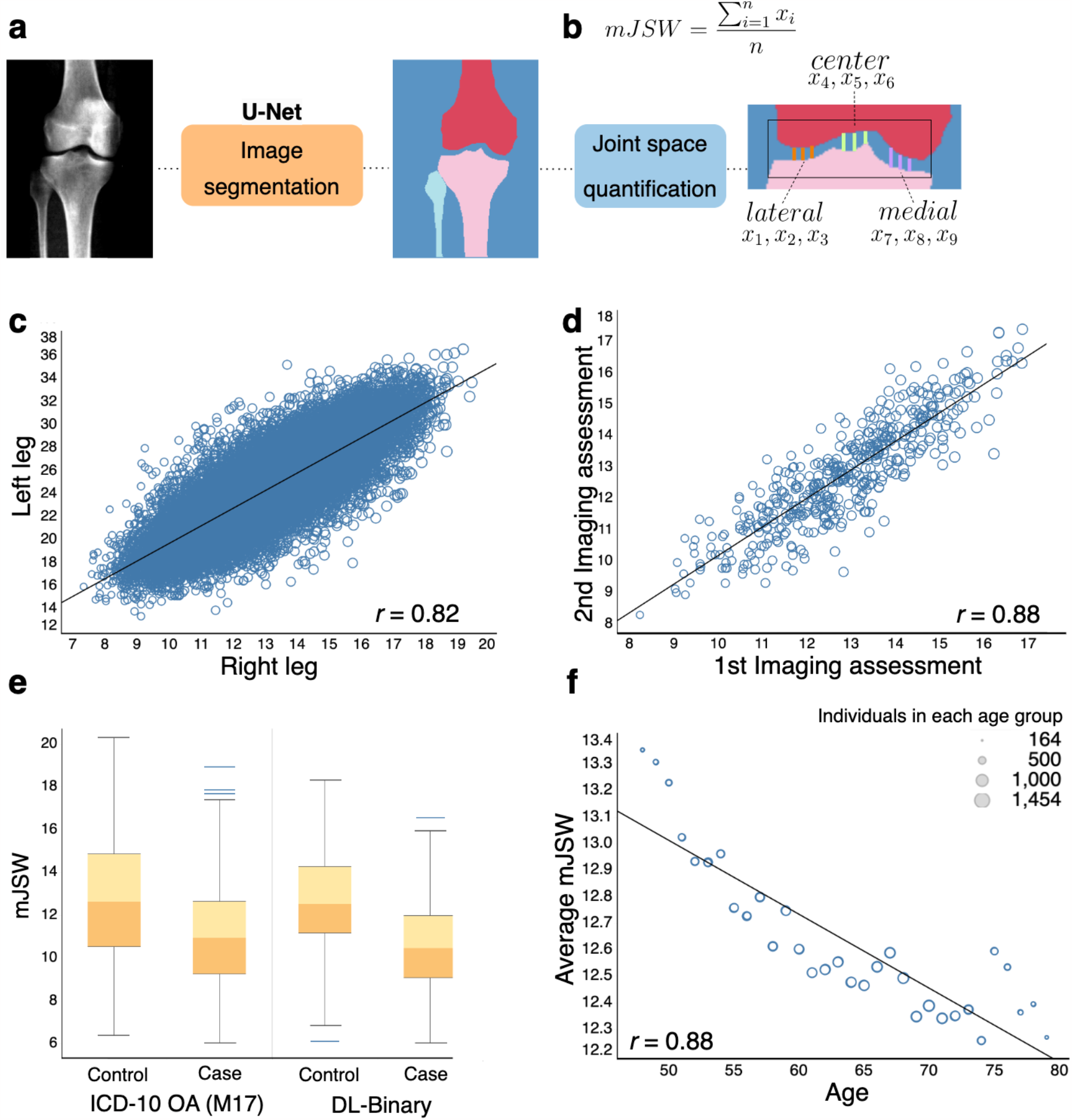
Deep learning based image segmentation for minimum joint space width (mJSW). **a** Deep learning based image segmentation labeling the femur, tibia, fibula, and background based on a U-Net architecture. **b** Measurement of mJSW between the tibia and the femur taken by the average of 3 points each in the lateral, center and medial portions of the knee joint. **c** Correlation of mJSW between the right and left leg of the same individual (n=29,257). **d** Correlation in calculated mJSW between the first and second imaging visit for the same individual (n=461). **e** mJSW is narrower in cases compared to controls, using both ICD-10 code M17 and DL-binary case identification. **f** Average mJSW decreases significantly with increasing age (ages 48 - 79, r = 0.88, p-value < 0.0001). Circle size corresponds to the number of individuals within each age group, with larger diameters equating to higher sample size relative to smaller circles.

We evaluated the performance of the segmentation model in several ways. First, the set accuracy, the correspondence between labeled data and annotation of the trained model on validation data, was 0.99. Second, the correlation between measurements taken between the right and left leg was 0.82 (**Fig. 2c**). Third, the correlation between images taken of the same person across two imaging visits was 0.88, despite changes in image resolution, scanner, technician and imaging position, demonstrating that our mJSW measurement process is fairly consistent across biological replicates (**Fig. 2d**). We do not expect to see 100% concordance across these replicates as joint space width often can change in a period of more than 2 years particularly in older individuals, in part due to possible knee joint cartilage degeneration. Fourth, we examined the relationship between mJSW and OA status, both using DL-binary and the ICD-10 code M17 case-control data (**Fig. 2e**). As expected, mJSW was significantly lower in cases compared to controls regardless of which case annotation we used (t-test p < 2.2 × 10^−16^, and p < 2.2 ×10^−16^ for DL-binary and ICD-10 code M17 respectively). Finally, we examined the relationships between mJSW and age - which is known to be highly associated with knee degeneration (**Fig. 2f**). Again, as expected, we found that the mJSW decreased significantly with age (linear regression, Beta = −0.028, p < 2.2 × 10^−16^).

### Genetic associations using image derived phenotypes

Having obtained IDPs related to knee OA, we performed GWAS to link these phenotypes to their genetic basis. After generating summary statistics for each genetic association (**Fig. 3**), we estimated SNP heritability using LD Score regression^24^ for the three phenotypes: (1) Knee OA as determined by the ICD-10 code M17 data from UKB, (2) Knee OA as determined using DL-binary and (3) mJSW, the quantitative phenotype highly correlated with severity of knee OA. The heritability of both binary phenotypes was low (ICD-10 code M17: 0.02 ± 0.02 and DL-binary: 0.04 ± 0.02). In contrast the heritability of the quantitative phenotype mJSW was 0.24 ± 0.02. Genomic inflation for the three phenotypes also confirmed this trend, with lambda for ICD-10 code M17: 1.0, DL-binary: 1.01, and mJSW: 1.06. Deviations from expectation across the genome are visualized in the qqplots inserts on **Fig. 3**. We found 18 independent loci that reached genome-wide significance in the mJSW GWAS, including one that was also significant in a previously reported GWAS for knee OA with 62,497 cases and 333,557 controls^25^ (**Fig. 3**). We found one locus and six genome-wide significant loci with either binary phenotype respectively (DL-binary and ICD-10 code M17), though mJSW and DL-binary had a genetic correlation of −0.92 ± 0.25 (**Methods**: Heritability and genetic correlation). This suggests substantial improvements in power from using a continuous, quantitative measure associated with knee OA disease severity.

**Fig. 3.**
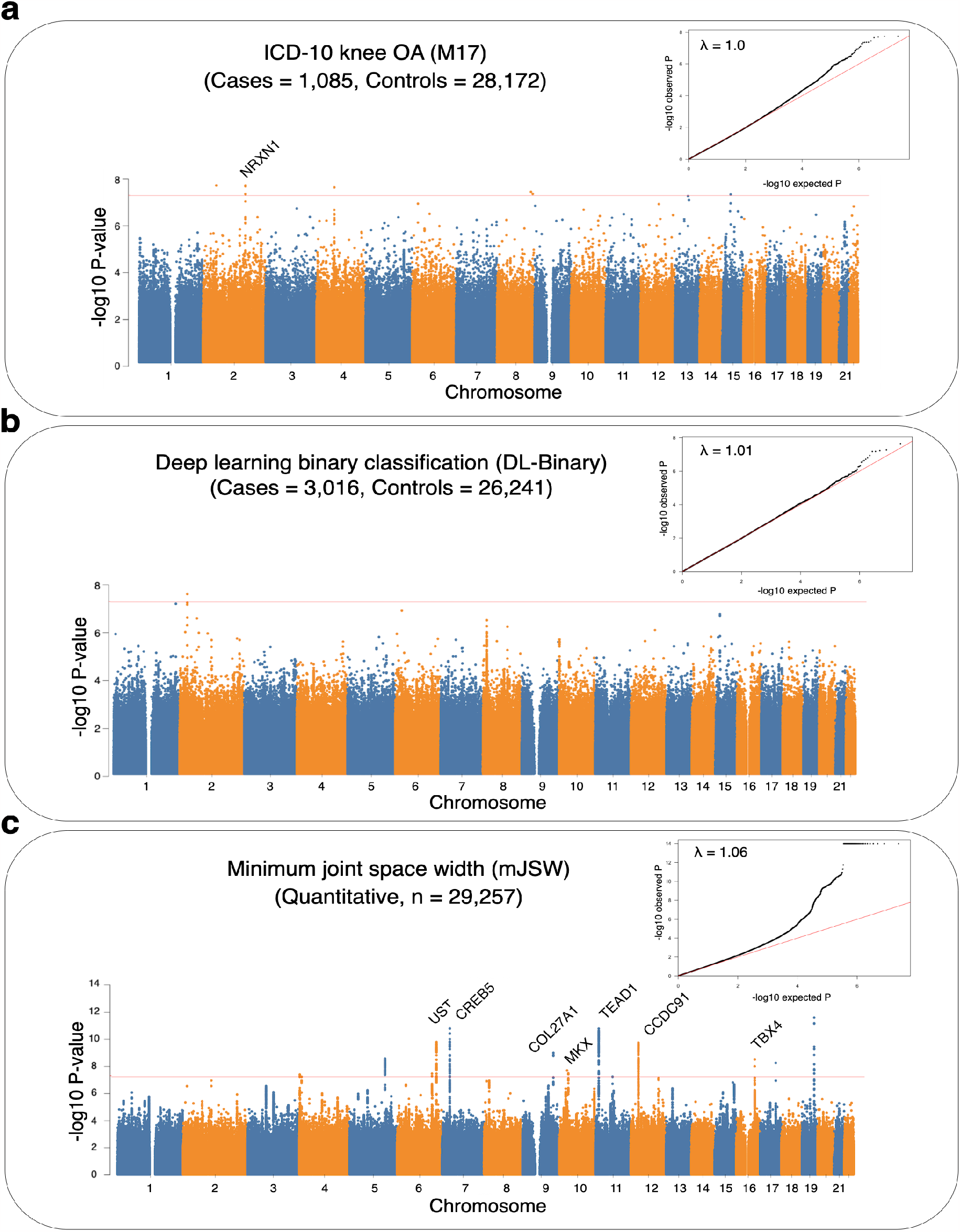
Manhattan plots for GWAS performed using three knee OA phenotyping methods. **a** ICD-10 code M17 defined knee OA case and control status. **b** The deep learning based automated case-control phenotype, DL-binary. **c** The deep learning based quantitative endophenotype, mJSW. Loci over the genome-wide significance threshold (p = 5 × 10^−8^) that are in close proximity to only a single gene are annotated. *Inset:* Quantile-quantile (qq) plot of deviation of the observed p-value from the theoretical distribution, along with the λ value quantifying genomic inflation.

### Polygenic risk scores for knee joint space are highly predictive of knee OA

As our GWAS for mJSW identified many more loci of genome-wide significance compared to DL-binary or ICD-10 code M17, we wanted to assess if this translated to improved power to predict knee OA in individuals outside of our DXA imaged sample. We computed PRS using clumping and thresholding (selecting variants below different p-value thresholds ranging from 1 to 1 × 10^−6^) from the GWAS of ICD-10 code M17, DL-binary and mJSW, and deployed these scores on 371,686 individuals in the population who were not included in the GWAS (**Methods:** Polygenic Risk Scoring). We carried out logistic regression with binary presence or absence of ICD-10 code M17 diagnosed knee OA as the outcome, using z-scores generated from each of the PRSs as the predictor variable, and the first 20 PCs, age, sex, height and BMI as covariates. After controlling for these variables and performing multiple hypothesis testing correction at the level of the total number of associations performed, the mJSW PRS remained independently associated with knee OA diagnosis regardless of p-value threshold, but the DL-binary PRS or the ICD-10 code M17 PRS were only significantly associated with knee OA at certain thresholds, again reflecting differences in power between the various GWASs (**Fig. 4a**).

**Fig 4.**
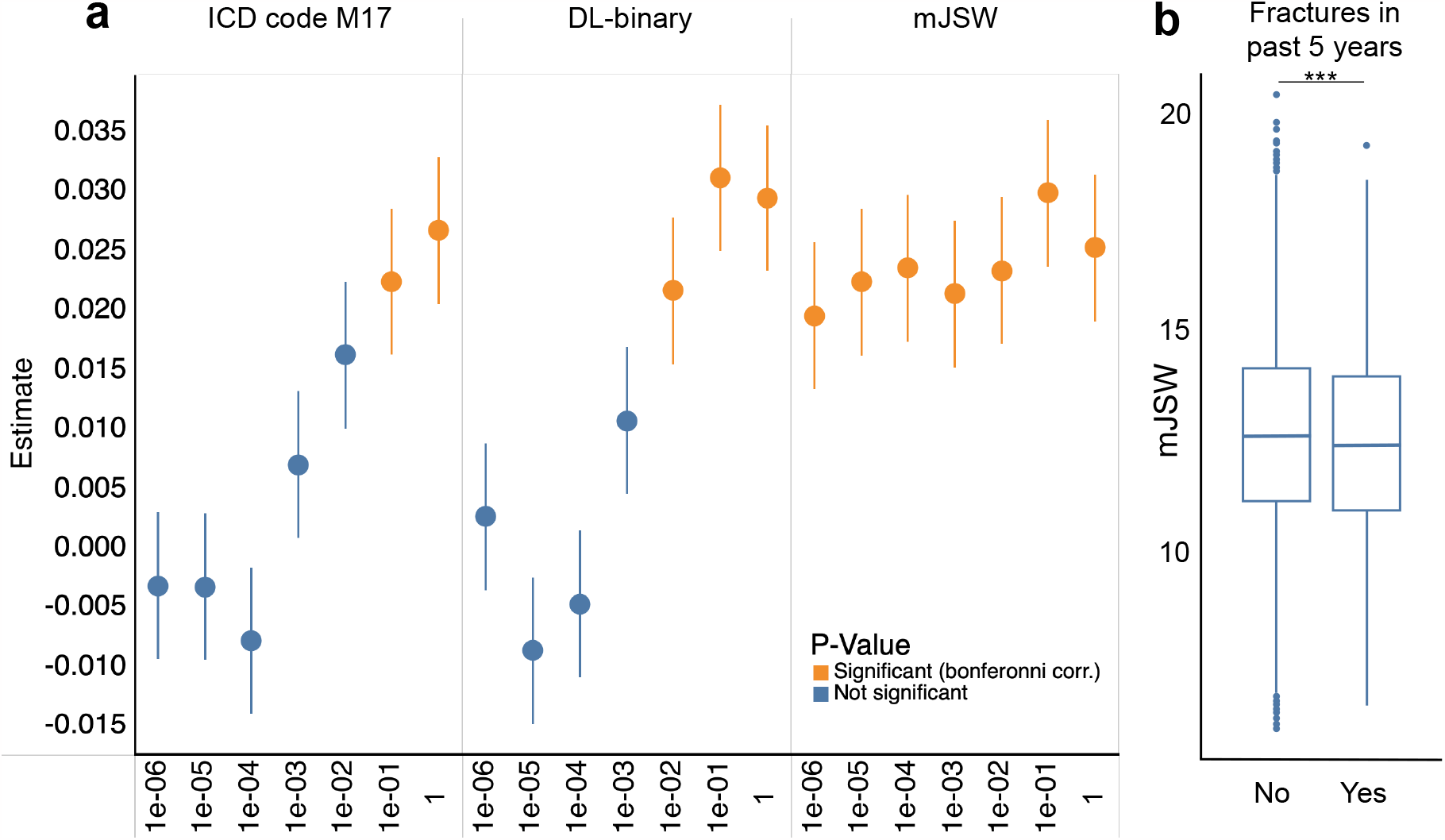
Genetic and epidemiological analysis of image derived phenotypes. **a** Results of performing logistic regression analysis using the PRS generated from each GWAS to predict ICD-10 M17 diagnosis on a hold-out dataset of 371,686 UKB individuals, showing the regression estimate obtained at each p-value threshold (and 1 standard error), colored by whether the test was significant after Bonferroni correction. **b** Boxplots showing the distribution of mJSW in patients who experienced a fracture in the past 5 years (n=24,147). mJSW was significantly predictive of fractures in logistic regression analysis, asterisks correspond to p-value significance (p < 1.10 × 10^−3^).

### Quantitative phenotyping allows for novel epidemiological associations

In addition to improving power in genetic analysis, we wanted to examine if we could use the mJSW phenotype to improve statistical power to detect an important epidemiological outcome in the health record, fractures within the last five years. After controlling for height, sex, age and body fat percentage, mJSW was significantly predictive of fracture in the last five years (p = 1.10 × 10^−3^) in logistic regression analysis, but not with DL-binary (fractures: p = 0.79) or with ICD-10 code M17 (p = 0.171) (**Fig. 4b**). While previous work on a much smaller sample size of ∼2000 individuals has shown that knee OA is associated with falls^26^, our results specifically implicate joint space narrowing with an independent increased risk of fractures, a known cause of death in individuals 65 and over^27^. These results emerge only upon examining our quantitative phenotype mJSW which captures an element of disease severity, revealing knee OA as an important risk factor for potentially fatal complications from fractures in older adults.

## Discussion

In this study, we demonstrate a deep learning method to directly phenotype OA cases and controls (DL-binary), as well as joint space narrowing (mJSW), from DXA scan derived AP view knee radiographs of the UKB. We compared this image derived phenotyping approach with case-control status already available in the ICD-10 record code M17 on the same set of individuals, to determine if image-derived phenotyping approaches have an effect on statistical power in GWAS.

We find that the case-control phenotyping using the DL-binary classification method enables us to raise the case count by greater than two fold and circumvents some issues with sourcing cases from the EHR such as variation in specific definitions of OA or differences in a clinician’s perception of the disease^28^. While previous work has shown that the ICD-10 record can have issues identifying individuals with disease for a variety of reasons, our study carrying out image based diagnosis at large scale provides evidence of the extent to which the record can be incomplete.

Additionally, both case-control methods lack information about disease severity, which may explain why they are underpowered compared to the quantitative measurement mJSW in the genetic and epidemiological analyses we investigate. The high genetic correlation between mJSW and DL-binary (92%) suggests that while the binary case-control phenotype of knee OA is underpowered compared with mJSW, the genetic relationships found between the two phenotypes are consistent with one another.

While computer vision approaches to extract and analyze DXA scan derived phenotypes are not themselves novel^16,17,29,30^, this work is amongst the first to use this approach on a disease for which diagnosis is primarily radiographic, to demonstrate that having a quantitative endophenotype that captures additional information about variation in disease severity improves power for genomic and epidemiological analysis. Although not based on the imaging data, two novel phenotyping methods leveraging deep learning to impute missing data in the UKB and to generate disease liability scores from binary case-control data in the EHR have shown significant boosts in statistical power for genomic studies^31,32^. Broadly, these and other approaches suggest that analysis of biobank data could benefit from quantitative refinement of disease phenotypes using alternative approaches.

One potential limitation of our study is that knee joint space narrowing is both causal and symptomatic in knee OA progression. As arthritis progresses, the joint space narrows due to the breakdown of cartilage, causing a resulting increase in pain and difficulty with movement. This narrowing of the joint space can also cause further damage, due to increased contact pressure at the affected joint. This makes it difficult to understand the root cause of knee OA with respect to the mJSW endophenotype, because joint space narrowing can both be a result of OA and a contributing cause to the progression of the condition. While the DL-binary method discovered two-fold more cases than what is annotated in the ICD-10 record, it is still likely to be an undercount due to our choice to use a particular instantiation of the model to limit the false positive rate as much as possible (**Fig. 1e**,**f**). Thus, despite improving the case-control ratio in the dataset, there may still be additional cases undetected by either method which could further improve statistical power in GWAS. Third, all GWAS in this work were restricted to individuals with European ancestry. Thus, the transferability of the specific findings in this genetic analysis (i.e. loci discovered from mJSW GWAS, trait heritability, and genetic correlation) across ancestries is not warranted without follow-up analyses.

Taken together, our study provides a proof-of-concept for the utility of quantitative phenotyping in biobank scale settings where a direct measurement of disease severity for a complex disease phenotype is possible. The results of this work suggest that this concept extends not only to other musculoskeletal diseases in which radiography is one of the primary methods for diagnosis (for example, directly measuring spinal curvature as opposed to scoliosis diagnosis), but to other analyses in which one can derive a quantitative alternative to case-control disease phenotyping.

## Methods

### UKB participants and dataset

All analyses were conducted with data from the UKB unless otherwise stated. The UKB is a richly phenotyped, prospective, population-based cohort that recruited 500,000 individuals aged 40–69 (mean 58) in the UK via mailers from 2006 to 2010^1^ (https://www.nature.com/articles/s41586-018-0579-z). In total, we analyzed data from 402,000 participants with genetic data of self-identified white British ancestry who had not withdrawn consent as of February 22, 2022. Of this genotyped cohort, 42,284 had available DXA imaging data. Access was provided under application number 65439.

### Dual-energy X-ray Absorptiometry (DXA) Imaging

The UKB has released DXA imaging data for a total of 50,000 participants as part of a bulk data field ID. The DXA images were collected using a Lunar iDXA instrument^1^ (GE healthcare) in DICOM format. A series of 8 images were taken for each patient: two whole body images - one of the skeleton and one of the adipose tissue, the lumbar spine, the lateral spine from L4 to T4, each knee, and each hip. Dual-energy X-ray absorptiometry (DXA) images were downloaded from the UKB bulk data. The bulk download resulted in 42,284 zip files, each corresponding to a specific identifier otherwise known as each subject’s EID. The uncompressed directories corresponding to each imaged subject contained several DXA images of the individual as described above. For this analysis, only images of the right and left knees from the AP view were used. It is important to note that all subjects in this analysis were instructed to lay flat on the DXA scanner machine during imaging, so that all resulting images are non-weight bearing.

### Phenotype and clinical data acquisition

The binary classification of patient disease phenotypes was obtained from a combination of primary and secondary ICD-10 codes. ICD-10 codes were truncated to only be the initial three characters. Patients received a “one” if a disease code appeared in their hospital records, and a “zero” otherwise. Reports of a fracture within the last 5 years of any visit (instance 0 to 3) was considered a case. Our classification of fractures increases case counts while excluding any childhood incidence.

### Computing infrastructure

We carried out all training using the Python programming language (www.python.org, version 3.7.7) with the PyTorch^33^ and Fastai version 1^34^ (https://github.com/fastai/fastai1) libraries on NVIDIA 1080-TI GPUs on the Maverick2 system and NVIDIA Quadro RTX 5000 GPUs on the Frontera system of the Texas Advanced Computing Cluster using the CUDA 11.1 toolkit.

### DXA scan image quality control and standardization

DXA images in DICOM format were first organized by anatomy following the manifest files located in each directory output by the imaging machine. DXA scans were subject to further quality control following the methods described in Kun et al., 2022^15^. Following initial data cleaning, AP view knee DXA scans were converted from DICOM to JPG format using the pydicom library^35^. To prepare a uniform set of images for segmentation, the numpy^36^ and opencv-python^37^ libraries was used to pad images to a standard width and height (800 ×1000 pixels), and outlier images that had resolutions outside of this standardized range were removed from all downstream analyses. Padded images were subject to further image resizing during training of the U-net architecture^23^ for segmentation (using a progressive resizing technique), but not during training of the classification model.

### Binary classification: DXA scan annotation procedure

546 images were sampled from the UKB for orthopedic surgeons to annotate. Images were sampled with reference to the ICD-10 code M17 to create a balanced dataset for training and validation. The KL grade^21^ based phenotype (DL-binary) was defined taking the following observations as input: presence or absence of osteophytosis, visible sclerosis of bone, and narrowing of the inter-bone joint space between the femur and tibia). Participating surgeons were instructed to annotate images as 0 or 1 based on whether or not each image qualified as KL grade 3 or greater, meaning that based on the radiographic evidence of knee OA the individual would be a candidate for joint replacement surgery. We considered 0 to be a control (but not necessarily devoid of any radiographic OA symptoms) and 1 to be a case (KL grade 3 or 4) warranting joint replacement. Surgeons went through a series of two rounds of independent grading on two datasets. Following a review of inter-rater reliability, the three annotating physicians went through a series of consensus grading, resulting in the final DL-binary dataset used for training and validation. For the DL-binary classification model, 80% of the data was reserved for training and 20% was reserved for validation.

### Binary classification: Normalization and data augmentation

Prior to performing binary classification, images were scaled to 224 × 224 pixels and normalized using ImageNet statistics. The ResNet-101 convolutional neural network (CNN) weights were initialized using the Kaiming normal method^22^. While training, multiple transformations were applied to the input images to regularize the model. These included a padding process as described above, as well as other transformations such as vertical flipping of the image, random rotation, zooming, warping, light and contrast change. This data augmentation was performed to improve the model’s ability to generalize in its predictions relative to variation in contrast and other image artifacts common to DXA scanning^38^.

### Binary classification: Network architecture and model training

We constructed a ResNet-101 CNN^22^ for our binary DXA image classifier, implementing transfer learning to reduce the amount of training time and resources for our classification task, using a pre-trained model obtained from training on the ImageNet^39^ (image-net.org) dataset and transferring the weights from this model to earlier layers of the network. We applied batch normalization and ReLU after each layer of the CNN to reduce overfitting and provide additional regularization using the Fastai version 1^34^ and PyTorch^33^ default parameters, and dropout was applied to the fully connected portion of the network. The output of the model is a binary classification for each DXA scan derived image passed in, a one-dimensional tensor containing values of 0 or 1 (control and case status), produced from passing the final layer of the network (the classification head) through the sigmoid and argmax activation functions. The batch size for all models was 64. We first plotted cross entropy loss as a function of learning rate in order to select the optimal hyperparameters. We trained the model for 42 epochs with discriminative learning rates ranging from 1 × 10^−3^ to 1 × 10^−6^.

### Image segmentation: DXA scan annotation procedure

We collected human generated annotations of each anatomical structure present in 63 DXA scans of the knee (40 training, 23 validation). Annotations were produced at the pixel level for each of the following segments of an AP knee DXA scan the: (1) femur, (2) tibia and (3) fibula. All annotations were reviewed by an orthopedic surgeon prior to training.

### Image segmentation: Network architecture and model training

We trained a U-net architecture^23^ with a 34-layer ResNet encoder^22^ to perform semantic segmentation of the knee joint, annotating the femur, tibia, and fibula coded as 1, 2 and 3 respectively at pixel-level resolution. We used a batch size of 4 for the segmentation model. We used the same transfer learning approach with the ImageNet dataset as described for the binary classifier, as well as a progressive upsampling strategy during training. First we down sampled masks to half their size, trained for 28 epochs, saved the model, then restarted the kernel and trained the saved model on regular now upsampled mask. This training procedure was used to efficiently utilize memory and reduce the model’s time to convergence. As described previously, we plotted cross entropy loss as a function of learning rate in order to select the optimal hyperparameters.

### Image segmentation: Measurement and quality control

After performing segmentation, we computed the minimum inter-bone knee joint space distance in pixels (of either leg), abbreviated as mJSW. Segmentation masks were processed using software developed for this analysis, written in python using the numpy^36^ and opencv-python^37^ libraries (https://github.com/briannaflynn/UKB_knee_segmentation). Labeled polygons within each segmentation mask were processed independently, converted to an identity matrix of ones and zeros (ones being the polygon processed, for example the femur, tibia or fibula). From this identity matrix, two matrices were produced from indexes produced and along the x and y axes. These indexes were used in the computation of basic features of the polygon such as maximum width, and maximum height. Indices were saved from this process and were later used to compute measurements of joint space width between the femur and tibia.

A major issue in combining our analysis across input pixel ratios was that these pixel ratios represented different resolution scalings due to variable distance between the scanner and the patient as a function of DXA scanner type and the size of the patient. To control for this scaling issue and to standardize the images, we chose to regress our mJSW measurements across all image resolutions with height obtained from the UKB. The estimates obtained from this regression were used to obtain a scaling factor for each image resolution that were then used for measurement normalization. We validated this regression and normalization procedure by comparing measurements taken on individuals who had DXA scans taken at two imaging assessments at different resolutions.

### Genetic QC

For all genome-wide association analyses, we filtered the participants to Caucasian individuals (FID 22006) from the white, British population as determined by genetic PCA (FID 21000) and participant surveys. We removed individuals whose reported sex (FID 31) did not match genetic sex (FID 22001), had evidence of aneuploidy on the sex chromosomes (FID 222019), were outliers of heterozygosity or genotype missingness rates as determined by UKB quality control of sample processing and preparation of DNA for genotyping (FID 22027), or had more than nine third-degree relatives or any of unknown kinship (FID 220021). In total 402,233 individuals remained. We further filtered to imaged participants (FID 20158) with complete DXA measurements (FID 12254); 33,475 remained.

Imputed genetic data for 487,253 individuals was downloaded from UKB for chromosomes 1 through 22 (FID 22828) then filtered to the quality-controlled subset using PLINK2^40^. All duplicate single nucleotide polymorphisms (SNPs) were excluded (--rm-dup ‘exclude-all’) and restricted to only biallelic sites (--snps-only ‘just-acgt’) with a maximum of 2 alleles (--max-alleles 2), a minor allele frequency of 0.1% (--maf 0.001), an individual missingness rates no more than 2.5% (FID 22005), and genotype missingness of no more than 5% (--maxMissingPerSnp 0.05). In total 14,846,570 SNPs remained in the imputed dataset. Non-imputed genetic data did not contain duplicate or multiallelic SNPs but were filtered to the quality-controlled subset; 703,993 SNPs remained.

### GWAS

GWAS was carried out using PLINK2, with a minor allele frequency of 0.001, a missingness per SNP of 5%, and a missingness per individual of 2.5%. Covariates were the first 20 genetic principal components provided by UKB (FID 22009), sex (FID 31), age (FID 21022), BMI (FID 21001) and standing height taken at the imaging assessment, instance 2 (FID 50). The final population size for all GWAS after both genetic and imaging QC was 29,257, and all GWASs had the same number of SNPs: 12,129,706. SNPs in each resulting GWAS were clumped using --clump with a significance threshold of 5.0 × 10^−8^, a secondary significance threshold of 1.0 × 10^−4^ for clumped SNPs, an r^2^ threshold of 0.1, and a 250 kb threshold of physical distance. SNPs were assigned to genes with --clump-verbose --clump-range glist-hg19.

### Heritability and genetic correlation

LD Score v1.0.1 was used to compute linkage disequilibrium regression scores per chromosome with a window size of 1 cM^24^ with the non-imputed genetic data. The heritability of each phenotype was then assessed using LD score regression^24^ with the same covariates as the GWAS. We examined the pairwise genetic correlation of the DL-binary and mJSW using GCTA version 1.93.2 beta for Linux^41^. We created the genetic relationship matrix for our quality-controlled subset with a minor allele frequency of 0.001, and then ran GCTA, using the first 20 genetic principal components provided by UKB (FID 22009), sex, age, BMI and standing height as covariates.

### Polygenic Risk Scoring

PRSs were computed with the IDP GWAS summary statistics in PLINK (v1.9) using the clumping and thresholding method. GWAS were clumped using an r^2^ threshold of 0.1 and a 250 kb threshold of physical distance for clumping. Significance thresholds of 1, 0.1, 1 × 10^−2^, 1 × 10^−3^, 1 × 10^−4^, 1 × 10^−5^, and 1 × 10^−6^ were used to compute PRSs for all three phenotypes run in GWAS. We then regressed ICD-10 code diagnosis of knee OA on the z-scores generated from each PRS obtained for each phenotype in all genotyped non-imaged individuals of white British ancestry (who had also undergone genetic QC), n = 371,723. In our logistic regressions we also controlled for age, sex, height, BMI and the first 20 principal components as covariates for all phenotypes.

### Transcriptome Analysis

To connect the genetics of joint space with the biology underlying synovial membrane differences in individuals with knee OA, as synovial fluid is functionally important in OA progression and inflammation^42,43^. We looked for enrichment of genes associated with our mJSW GWAS in gene expression data obtained from synovial tissues in 12 knee OA patients^44^. A set of inflamed as well normal synovial tissue was obtained from each patient for a set of 24 data points. This microarray gene expression data was obtained from the GEO data repository GSE46750, and the data were quantile normalized and log-transformed. Gene level p-values for our skeletal phenotype GWAS were first calculated using the positional mapping tool with default settings in SNP2GENE (version 1.3.7)^45^. We then performed gene property analysis in MAGMA (version 1.10)^46^ to determine associations between genes implicated in our mJSW GWAS and genes expressed in normal as well as inflamed synovial tissue. We found enrichment in our mJSW genes for gene expression from both normal and synovial tissue obtained from knee OA patients but no difference in enrichment from differential expression of normal and inflamed tissue **Table 1**.

**Table 1.** Results of enrichment analysis for genes significantly associated with the mJSW phenotype from gene expression data obtained from normal (normal_avg) and inflamed (inflammatory_avg) synovial tissue. The statistics are produced from a one-sided association test between the phenotype and the 12 normal and 12 inflammatory data points.

| VARIABLE | ESTIMATE | ESTIMATE_STD | SE | P-VALUE |
| --- | --- | --- | --- | --- |
| normal_avg | 0.017 | 0.029 | 0.004 | 2.84E-05 |
| inflammatory_avg | 0.017 | 0.029 | 0.004 | 3.05E-05 |

## Data Availability

All our data comes from the UK Biobank Study under application ID 65439.

## Data availability

Deep learning and image processing tools can be found at tools can be found at https://github.com/briannaflynn/UKB_knee_segmentation and https://github.com/briannaflynn/dxaconv/. GWAS summary statistics are available at https://utexas.box.com/s/8stbz74t9hrx7fdbgl0gcqi66miodl92. Individual level information of image derived phenotypes has been reported back to the UKB and will be available upon publication.

## Acknowledgements

This research has been conducted using the UKB Resource under Application Number 65439. We thank Olivia Smith for insightful discussions and comments regarding bioinformatic analysis.

V.M.N was supported on a grant from the Allen Discovery Center program, a Paul G. Allen Frontiers Group advised program of the Paul G. Allen Family Foundation and a Good Systems for Ethical AI grant from the University of Texas at Austin. B.F. was supported on an NSF Graduate Research Fellowship DGE 2137420. E.M.J. and B.F were supported by an NIH T32 grant 5T32LMO012414. B.F. was also supported on a UT Austin Provost’s Graduate Excellence Fellowship.

## Author contributions

B.F., E.J. and V. M. N. wrote the paper with input from all co-authors. B.F., E.J., E.K., A.G, K.K., K.A. and E.L. performed analysis. T.J., Z.T., P.J. and V.M.N. supervised the analysis.

